# Planning for the End at the Beginning: A Lesson in Sharing Research Findings of a Community-based COVID-19 Seroprevalence Survey

**DOI:** 10.1101/2023.11.27.22273609

**Authors:** Anne M. Trolard, Emily Hickner, Victoria V. Anwuri, Kimberly J. Johnson, Charles W. Goss, Rachel E. Cohen, Kate Donaldson, Elvin Geng, Brett B. Maricque

## Abstract

**Introduction:** Sharing research findings with participants is important but challenging. We evaluated a plan to share findings with participants of a COVID-19 seroprevalence project.

**Methods:** An electronic survey was distributed to participants after completion of the project to determine reach of planned coverage of the findings.

**Results:** Most respondents (80% n=428) had not seen the coverage; but nearly all (90%, n=388) wanted to see the findings. Participants identified a brief visual report as their preferred avenue. A second follow-up survey found that 99% of those who read the report approved.

**Discussion:** When researchers alone create a plan to share project findings, efforts may not reach nor be in the format desired by participants.

**Conclusions:** This work can serve as a model for collaborating with community to disseminate public health data.

## Introduction

People who participate in research studies consistently state that they would like to receive aggregate findings from studies in which they participate^1–4^. Participants report it is their right to hear about the findings^5^ and that sharing findings builds trust between researchers and communities. Conversely, trust erodes when researchers do not share findings. The collective benefits of sharing findings extend beyond a specific study and accrue when researchers commit to sharing^3^.

Health researchers do not routinely share findings with study participants even though they express a desire, and sometimes even an intention, to do so^6^. Researchers report many barriers to such sharing, including financial, time, and know-how constraints; as well as concerns about presenting findings to a lay or non-scientific audience^7^.

Researchers’ concerns regarding participant literacy are valid, and ensuring participants are not harmed by the findings is imperative. There is certainly not a one-size-fits-all approach. Without mandates to share, much less a framework or values system that prioritizes participants’ right to receive results, it is easy to see why researchers often fall short of their intentions. The COVID-19 pandemic has shown us that engaging people in the scientific process is crucial^8^.

We conducted a COVID-19 seroprevalence and survey project in St. Louis County between August and October 2020^9^, which included a plan to share aggregate project findings with participants and the entire community. The communication plan was put together by a team that included media professionals, academics, and community partners, but not participants. The goal of the present study was to assess the extent to which participants received and were satisfied with the presentation of the project findings.

## Materials and Methods

The project was conducted in collaboration with the St. Louis County Department of Public Health. A total of 2314 residents agreed to take a telephone survey that included questions about their Covid-19 experiences and behaviors, and of those 1435 provided an email address as part of registering for testing. Following conclusion of the project, findings were shared through standard media channels, including: 1) a press conference hosted by the county executive; 2) a media advisory posted on the county website; 3) an article in a local magazine; 4) a local radio interview with the Principal Investigator; 5) segments on local TV; and 6) articles by Washington University School of Medicine.

After the project, the authors drafted a brief follow-up survey to understand if participants saw coverage of the project findings, and if they found the coverage satisfactory. This survey also asked what specific information participants might like to receive and how they would like to receive it. The follow-up survey was sent by email to participants who provided an email address.

## Results

The follow-up survey was delivered to 1266 valid email addresses and of those 535 (42%) responded. This respondent group had 3% more females and 16% more white respondents compared to those who participated in the seroprevalence project^9^.

An overwhelming majority of respondents, 80% (n=428), had not seen the project findings. Those who had (n=107, 20%) were most likely to have read about the findings in a Washington University communication (n=45, 42%); seen coverage on TV (n=31, 29%); or read the media advisory on the county website (n=17, 16%). Ninety-nine percent of the 107 participants who had seen the findings were satisfied with the information that was shared.

Ninety percent of those who did not see the findings reported wanting to see findings (n=388). A total of 466 respondents shared the findings that most interested them. Figure 1 details these preferences, showing that they cared most about seeing project findings and how the county used the findings to help handle the pandemic. Most respondents wanted to hear about the findings in a 1-2 page visual report sent by email (n=343), followed by a 1-2 page visual report sent by mail (n=101) and attending a pre-recorded webinar discussing the findings (n=60).

**Fig 1.**
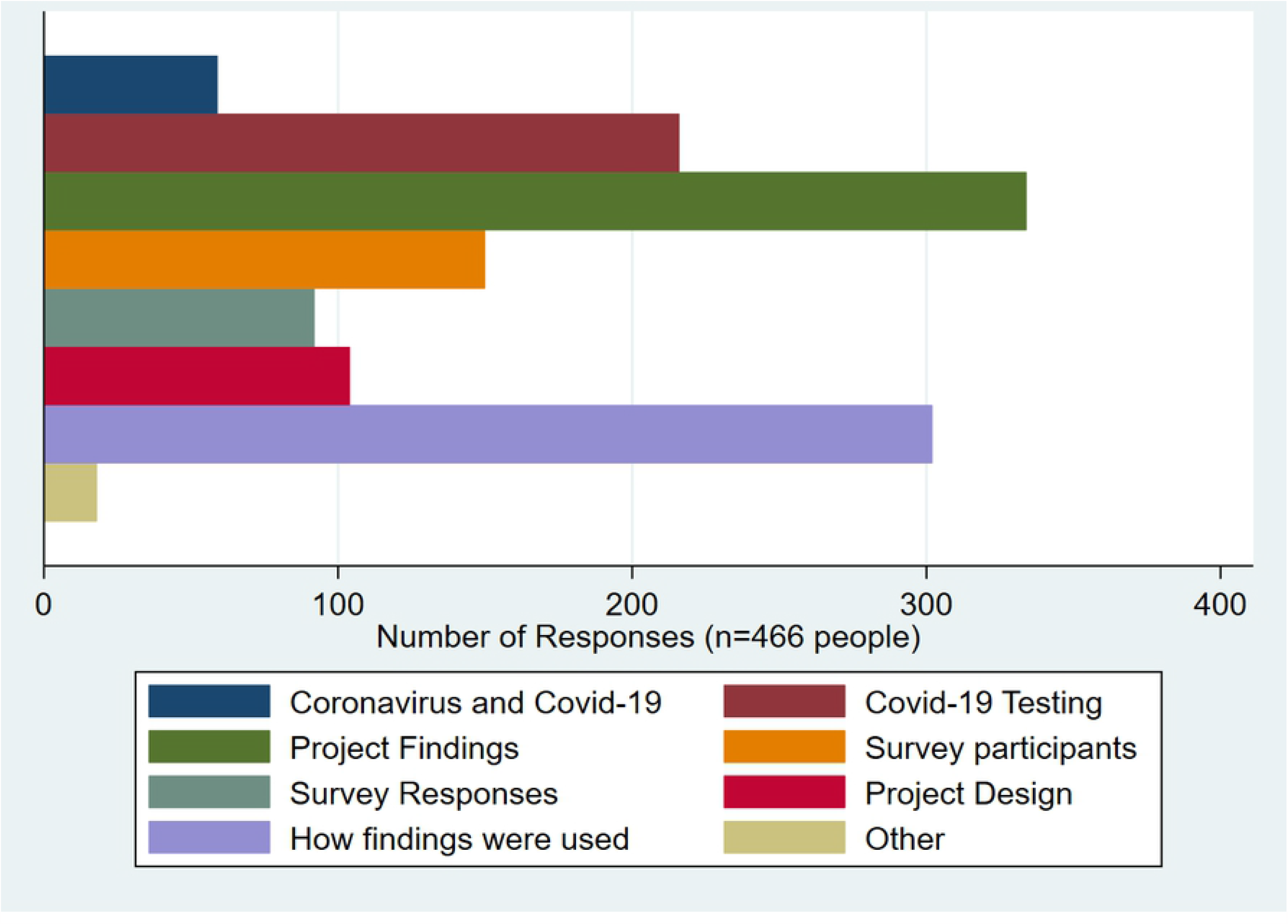
Specific information and findings that participants wanted shared. Respondents could select multiple answers to this question. Responses are presented in the order they were asked.

We drafted a visual report that included information requested by respondents and then mailed and emailed per respondent request. Four weeks later, we followed up with respondents to solicit feedback on the visual report and assess their interest in receiving future findings (e.g., publications). One-hundred thirty-eight people provided feedback (31%). Of the 102 that read the report, 99% indicated it was “Excellent” or “Pretty Good”. Additionally, 48% of respondents expressed interest in participating on a community review board to review public health projects.

## Discussion

Despite earnest efforts by our team to share the project findings using a diffusion strategy, our efforts did not reach most respondents. This suggests that a diffusion approach for sharing findings of public health projects can be ineffective in reaching the majority of participants, and dissemination planning is needed. Notably, respondents most commonly encountered findings in press releases from Washington University. We suspect that many of these individuals had prior contact with the University or sought out project findings themselves using the internet.

Respondents were willing to offer their preferences about how our team should share project findings. The vast majority preferred to receive findings in a digital visual report. This preference illustrates why project participants should be actively engaged in developing a plan for sharing findings. Dissemination strategies could be developed in collaboration with project participants prior to the end of the study, rather than solely by researchers. This early collaboration is what participants themselves suggest^5^ and could lead to projects where researchers engage participants as co-creators of new knowledge.

Our follow-up survey was limited to those for whom we had email addresses, thus a visual report may not be the preferred mode of communication for all participants. However, the purpose of this work was not to identify the single best mode of communication, but rather to demonstrate that engaging project participants in discussions about how to share findings is manageable and fruitful.

The follow-up survey was not planned from the project’s outset, and therefore the resources needed to carry it out were limited. To our surprise, the overall process was not resource intensive, yet we did find that specific resources were essential to our success. First, we were able to re-contact project participants using email addresses collected during enrollment. Second, we were able to use the database infrastructure established for the seroprevalence testing phase of the project. Third, and most importantly, our team included a graphic designer and a health communication and training specialist who had the skill set to produce the visual report. Our future work will build on this experience to engage participants in the planning and execution of public health research projects.

## Conclusions

The work reported here was well-received by participants and was rewarding for the research team. Our team received dozens of personal emails expressing support and appreciation for our commitment to sharing the project’s results. We learned directly from the people our work set out to serve, and established a foundation for engaging people in the scientific process. This work can serve as a model for collaborating with community members to improve dissemination of public health data. It is also a step in the right direction for building trust between research institutions and the public.

## Supporting information

IRB Determination Memo

## Data Availability

The data underlying the results presented in the study are available from the St. Louis County Department of Public Health.

## Acknowledgements

We thank the St. Louis County Department of Public Health for their support and partnership on this project. We thank the residents of St. Louis County for their contributions to the COVID-19 seroprevalence survey and their engagement in this project. This project was funded by the St. Louis County Department of Public Health.

