## Supplementary material for "Planning for the End at the Beginning: A Lesson in Sharing Research Findings of a Community-based COVID-19 Seroprevalence Survey": IRB Determination Memo

**IRB ID #:** 202204043

**To:** Anne Trolard

**From:** The Washington University in St. Louis Institutional Review Board,
WUSTL DHHS Federalwide Assurance #FWA00002284
BJH DHHS Federalwide Assurance #FWA00002281
SLCH DHHS Federalwide Assurance #FWA00002282

**Re:** Sharing Covid-19 Survey Results

**Human Subjects Research Determination:**

**Not HSR**

Because the purpose of the follow up survey is tied to the St. Louis County Department of Public Health project that was previously conducted and the information collected will not be used to answer a research question or to publish generalizable research results, this activity is not considered to meet federal definitions under the jurisdiction of an IRB and therefore falls outside the purview of the HRPO.

This determination has been electronically signed by IRB Chair or Chair Designee:
Jennifer Maynard, BA, Psychology
04/07/22 1809
